# Growth Differentiation Factor 15 Connects Stress and Ageing in a UK Biobank Cohort

**DOI:** 10.1101/2025.07.04.25330886

**Authors:** Steven Lehrer, Peter H. Rheinstein

## Abstract

**Background:** The brain–body energy-conservation model proposes that signals from metabolically active senescent cells, including growth differentiation factor 15 (GDF15), prompt the brain to downregulate systemic energy expenditure, leading to functional decline.

However, large-scale human studies testing this hypothesis are lacking. We examined whether circulating GDF15 is associated with physiological and cognitive ageing phenotypes and whether it mediates the relationship between psychological stress and muscle strength.

**Methods:** We analyzed data from the UK Biobank, a prospective cohort of over 500,000 adults. Circulating GDF15 concentrations were measured using Olink proteomics. Outcomes included insulin resistance (TG/HDL-C ratio), maximum heart rate during fitness testing, hand grip strength, C-reactive protein, telomere length, fluid intelligence, and physical activity. Psychological stress was assessed via self-report. Associations were evaluated using multivariate linear regression adjusting for age and sex. Mediation analysis tested whether GDF15 mediated the association between stress and grip strength.

**Results:** Higher GDF15 was significantly associated with reduced grip strength (β = –0.180, p < 0.001), higher insulin resistance, lower maximum heart rate, higher C-reactive protein, shorter telomere length, diminished fluid intelligence, and reduced physical activity (all p < 0.001). Mediation analysis demonstrated that GDF15 partially mediated the association between psychological stress and grip strength (indirect effect = –0.053, 95% CI –0.074 to –0.033).

**Conclusion:** These findings support the brain–body energy-conservation model, suggesting that GDF15 serves as a systemic signal linking psychological and cellular stress to declines in physiological and cognitive function. Targeting GDF15 pathways may represent a novel strategy to mitigate stress-related ageing processes.

**Graphical Abstract Caption:** Chronic psychological stress increases circulating GDF15, which signals the brain and contributes to systemic ageing phenotypes including reduced muscle strength, lower maximum heart rate, shorter telomere length, higher insulin resistance, and decreased physical activity. Mediation analysis in the UK Biobank (N = 500,000) demonstrated that GDF15 partially mediates the association between stress and reduced muscle strength, supporting the brain–body energy-conservation model.

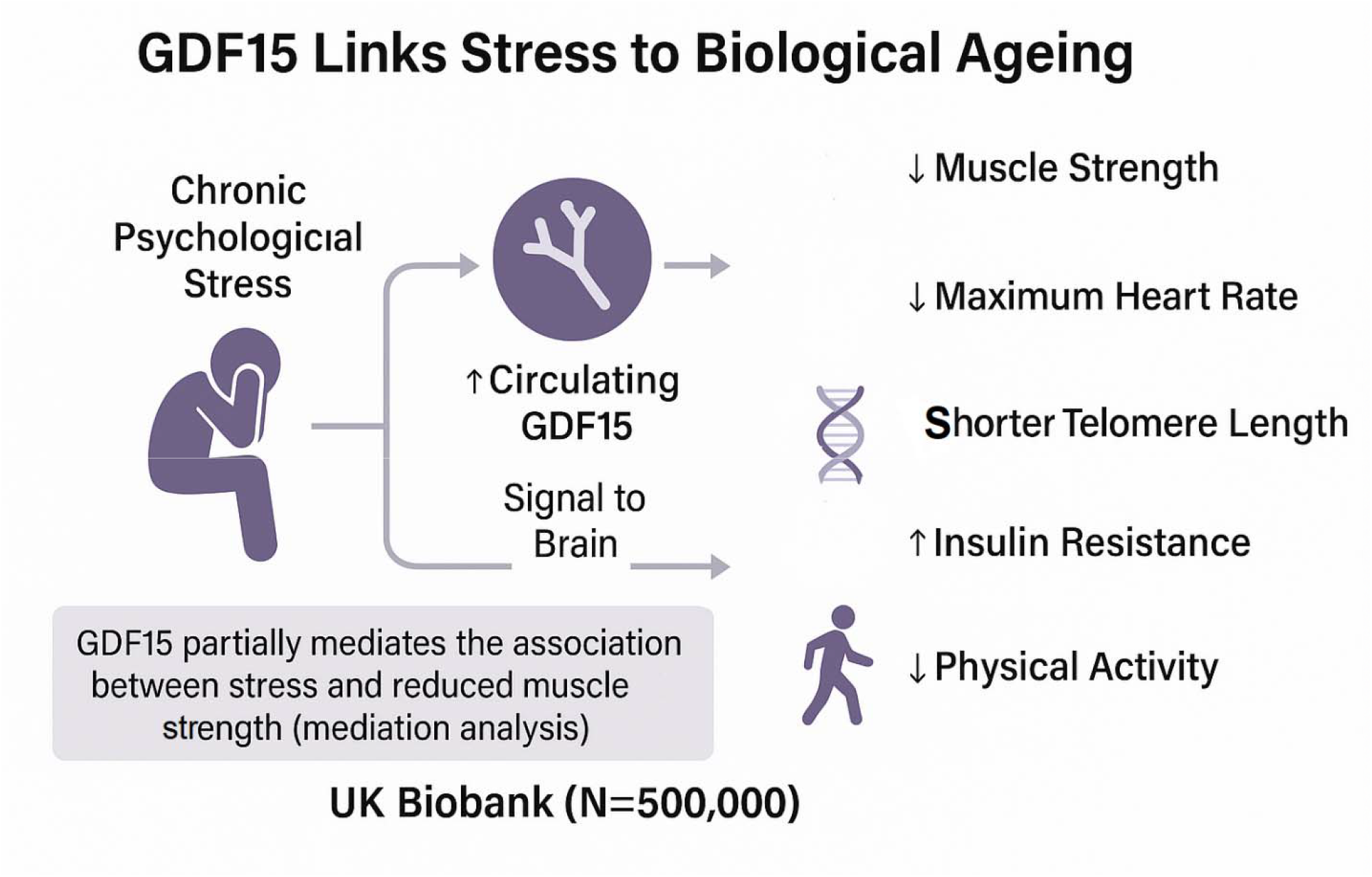

---

Ageing is increasingly understood as a systemic, multi-level process shaped by the interplay between cellular damage, metabolic adaptations, and central nervous system regulation. A growing body of evidence implicates cellular senescence, chronic inflammation, and psychological stress as key contributors to accelerated ageing phenotypes. Traditionally, senescent cells, that is, cells that have permanently exited the cell cycle, were thought to be metabolically quiescent. However, recent observations challenge this assumption [1]. Sturm et al. reported that senescent human skin cells exhibit a markedly elevated metabolic rate compared to proliferating cells, suggesting that senescence imposes a substantial energetic burden on the organism [2].

Metabolically active senescent cells secrete signals, inflammatory cytokines such as growth differentiation factor 15 (GDF15). Although GDF15 is produced by many organs, its receptor GFRAL is found only in the brain, suggesting that GDF15 is responsible for sending the brain signals about cellular stress. These signals communicate increased cellular energy demands to the brain. In response, the brain downregulates energy allocation to other physiological processes, leading to systemic features of ageing—such as reduced muscle mass (sarcopenia), greying hair, lower thyroid hormone production, and increased insulin resistance.

Psychological stress has long been recognized as a potent accelerator of biological ageing. Chronic stress activates the hypothalamic–pituitary–adrenal (HPA) axis and increases circulating glucocorticoids, which in turn contribute to inflammation, oxidative stress, and metabolic dysregulation. Large cohort studies have demonstrated that prolonged stress exposure is associated with shorter leukocyte telomere length, altered DNA methylation profiles, and increased risk of age-related diseases. Moreover, psychological stress has been linked to impaired muscle function, reduced cardiovascular capacity, and insulin resistance, all of which are hallmarks of declining physiological resilience. In the context of the brain–body energy-conservation model, stress may amplify signalling pathways, such as those involving GDF15, that inform the brain of elevated cellular energy demands, thereby accelerating compensatory downregulation of metabolic processes. Clarifying how psychological stress contributes to these pathways could provide critical insights into the mechanisms underlying stress-related vulnerability to ageing phenotypes [1].

Building on these findings, Picard and colleagues have advanced the *brain–body energy-conservation model*, a conceptual framework positing that the brain responds to signals from energetically costly senescent cells—mediated in part by cytokines such as GDF15—by downregulating other physiological processes to maintain energy balance [3]. This adaptive mechanism may help explain why local hypermetabolism at the cellular level coexists with global declines in energy expenditure and functional capacity in ageing organisms. Intriguingly, chronic psychosocial stress appears to exacerbate this downward trajectory, potentially accelerating telomere shortening, epigenetic ageing, and inflammation [4, 5].

Despite the conceptual appeal of the brain–body energy-conservation model, large-scale human data testing its core predictions remain limited. Specifically, it is not known whether biomarkers of cellular senescence and stress-related signalling are associated with systemic ageing outcomes in population cohorts. The UK Biobank (UKBB), a prospective cohort study of over 500,000 adults with deep phenotyping, linked health records, and genetic data, offers an unprecedented opportunity to test key elements of this hypothesis in vivo. For example, UK Biobank participants have longitudinal measures of psychological stress, circulating inflammatory markers, and physical health trajectories, alongside genomic and epigenetic data relevant to cellular ageing.

The present study leverages UK Biobank data to evaluate associations among chronic stress exposure, circulating GDF15 concentrations, and phenotypic indicators of ageing. By integrating psychosocial, biochemical, and health outcome data, we aimed to assess whether elevated GDF15 mediates the relationship between stress and ageing phenotypes, and test whether these associations are moderated by genetic and environmental factors. Clarifying these links has the potential to validate or refine the brain–body energy-conservation model and to identify targets for interventions designed to decouple stress responses from accelerated biological ageing.

## Methods

GDF15 was measured using Olink proteomics technology with the Olink Explore HT proteomics platform from Thermo Fisher Scientific, which employs the Proximity Extension Assay (PEA) technology coupled with Next-Generation Sequencing (NGS) readout [6]. This technology allows for highly multiplexed, specific, and sensitive measurement of thousands of proteins simultaneously from very small sample volumes (as low as 2 µL of plasma or serum), making it suitable for large-scale population studies like the UK Biobank.

Insulin resistance was assessed with an insulin resistance surrogate, the TG:HDL-C ratio described by Olivieri et al [7]. The ratio was calculated using non-fasting serum TG and HDL-C measurements (UK□Biobank fields 23400, 23401, 23407). Individuals with elevated TG/HDL-C have more insulin resistance.

In a subset of subjects (∼20,000) who had wearable electrocardiogram (ECG) monitoring, UKBB assessed heart rate variability, and episodic maximum elevated heart rates were captured (field 6033).

Hand grip strength, a proxy for muscle mass and sarcopenia, was measured using a standardized protocol at the baseline assessment visit. Participants were seated with their elbow flexed at 90 degrees and their upper arm resting against the torso.

Using a Jamar J00105 hydraulic hand dynamometer, they were instructed to squeeze the handle as hard as possible for approximately three seconds. Grip strength was recorded separately for the right and left hand in kilograms. If participants were unable or unwilling to perform the measurement with one hand, the other hand alone was tested. Data were captured as continuous variables (UK Biobank Field IDs 46 and 47). For analysis, the maximum grip strength recorded across both hands was used as the primary indicator of muscle strength. This approach is consistent with prior epidemiological studies and aligns with consensus definitions of low grip strength as a marker of probable sarcopenia.

The UK Biobank assesses fluid intelligence primarily through a dedicated 13-question verbal-numerical reasoning test administered under a two-minute time limit. While brief and custom-made, it serves as a valuable measure within this large-scale epidemiological resource [8].

Leukocyte telomere length was assessed by UK Biobank from baseline blood samples using a validated quantitative polymerase chain reaction (qPCR) assay. The assay measured the ratio of mean telomere repeat copy number to a single-copy gene (T/S ratio), which was then log-transformed and standardized. Details of the laboratory protocol, quality control procedures, and assay performance have been described previously [9]. In brief, DNA was extracted from leukocytes and analyzed in triplicate, with rigorous inter-plate and intra-plate calibration to minimize assay variability. The resulting T/S ratio reflects relative telomere length, with higher values indicating longer telomeres. For analysis, the z-standardized telomere length (Field ID **22191**) was used as a continuous variable, with higher values reflecting longer telomeres relative to the sample mean. Samples with missing or unreliable telomere length measurements were excluded.

Psychological stress was assessed using UK Biobank Data-Field **6145**, which records participants’ self-reported experience of illness, injury, or bereavement stress in the past two years. Specifically, respondents were asked: *“In the last two years, have you experienced any periods of stress that have lasted for more than one month?”* Response options included *“Yes”, “No”, “Do not know”*, and *“Prefer not to answer.”* For the present analysis, stress was defined as a binary variable: participants responding *“Yes”* were classified as having experienced prolonged stress, while those responding *“No”* were classified as not experiencing prolonged stress. Responses of *“Do not know”* and *“Prefer not to answer”* were treated as missing data and excluded from analysis.

We conducted mediation analysis to examine whether circulating GDF15 levels mediated the relationship between psychological stress and right-hand grip strength. The analysis was performed using the PROCESS macro (version 5.0) for SPSS [10], specifying Model 4, which estimates direct and indirect effects of an independent variable (stress) on a dependent variable (grip strength) through a single mediator (GDF15). Age and sex were included as covariates in all models to control for potential confounding. Bootstrapping with 5,000 resamples was used to generate bias-corrected 95% confidence intervals for the indirect effect. An indirect (mediated) effect was considered statistically significant if the bootstrap confidence interval did not include zero. Standardized and unstandardized coefficients were reported for all paths.

## Results

Figure 1 shows GDF15 versus maximum heart rate on electrocardiogram during fitness test, 3,807 cases. The inverse correlation is significant (p < 0.001).

**Figure 1.**
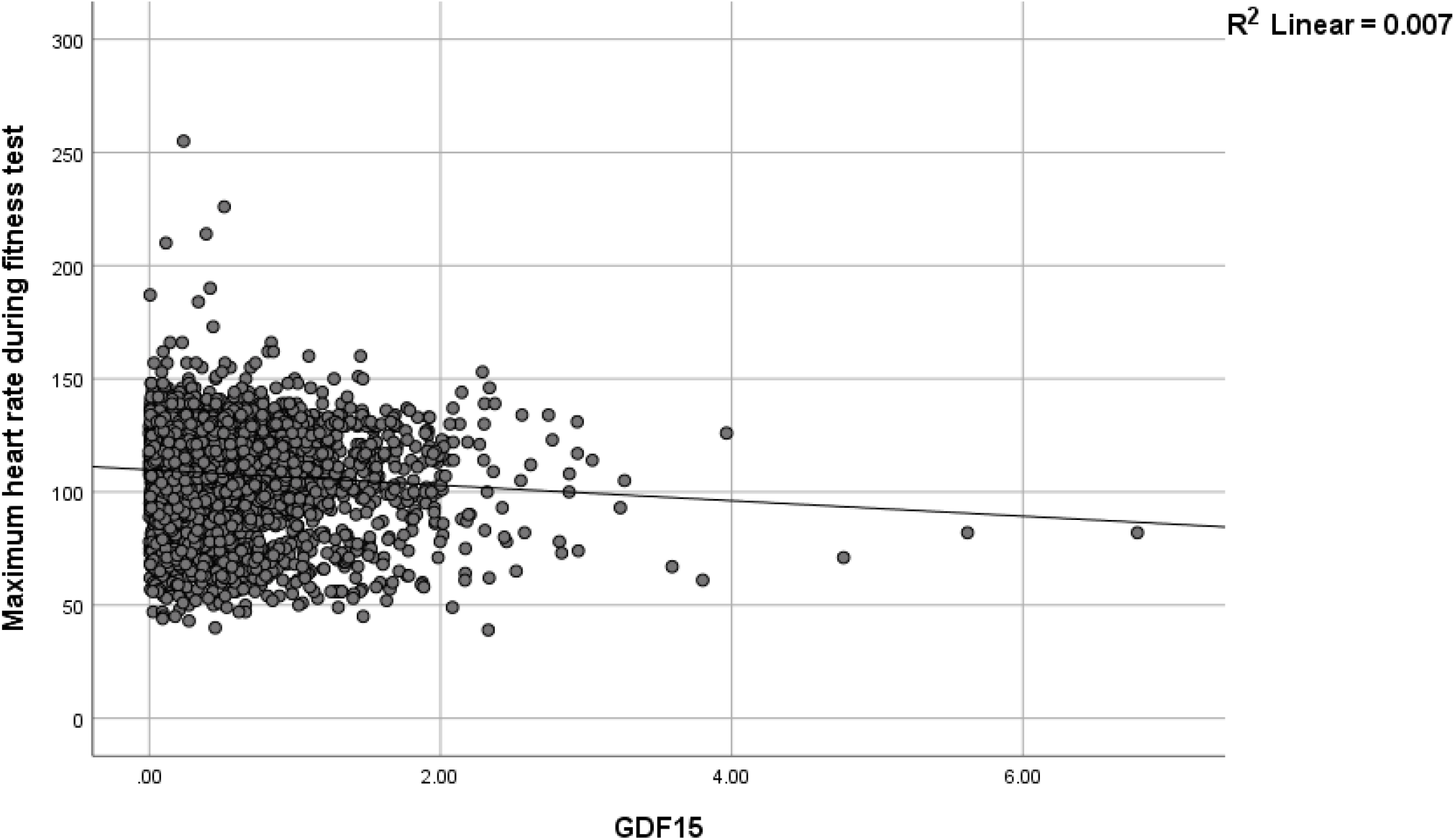
GDF15 versus maximum heart rate during fitness test, 3,807 cases. The correlation is significant (p < 0.001).

Multivariate linear regression was conducted to examine the association of age, sex, and circulating GDF15 levels with maximum heart rate during fitness test. The overall model was statistically significant p < 0.001, accounting for approximately 3.4% of the variance in heart rate. Higher GDF15 levels were significantly associated with a lower heart rate, β = – 0.052, p = 0.001). Male sex was associated with a modestly lower heart rate compared to female sex (β = – 0.045, p < 0.001). Additionally, older age predicted lower heart rate (β = – 0.15, p < 0.001). These results indicate that circulating GDF15, male sex, and higher age are each independently associated with reduced maximum heart rate during fitness testing in this cohort.

Figure 2 shows GDF15 versus insulin resistance (triglycerides/HDL-C, 29,811 cases). The correlation is significant (p < 0.001). Individuals with elevated GDF15 have more insulin resistance.

**Figure 2.**
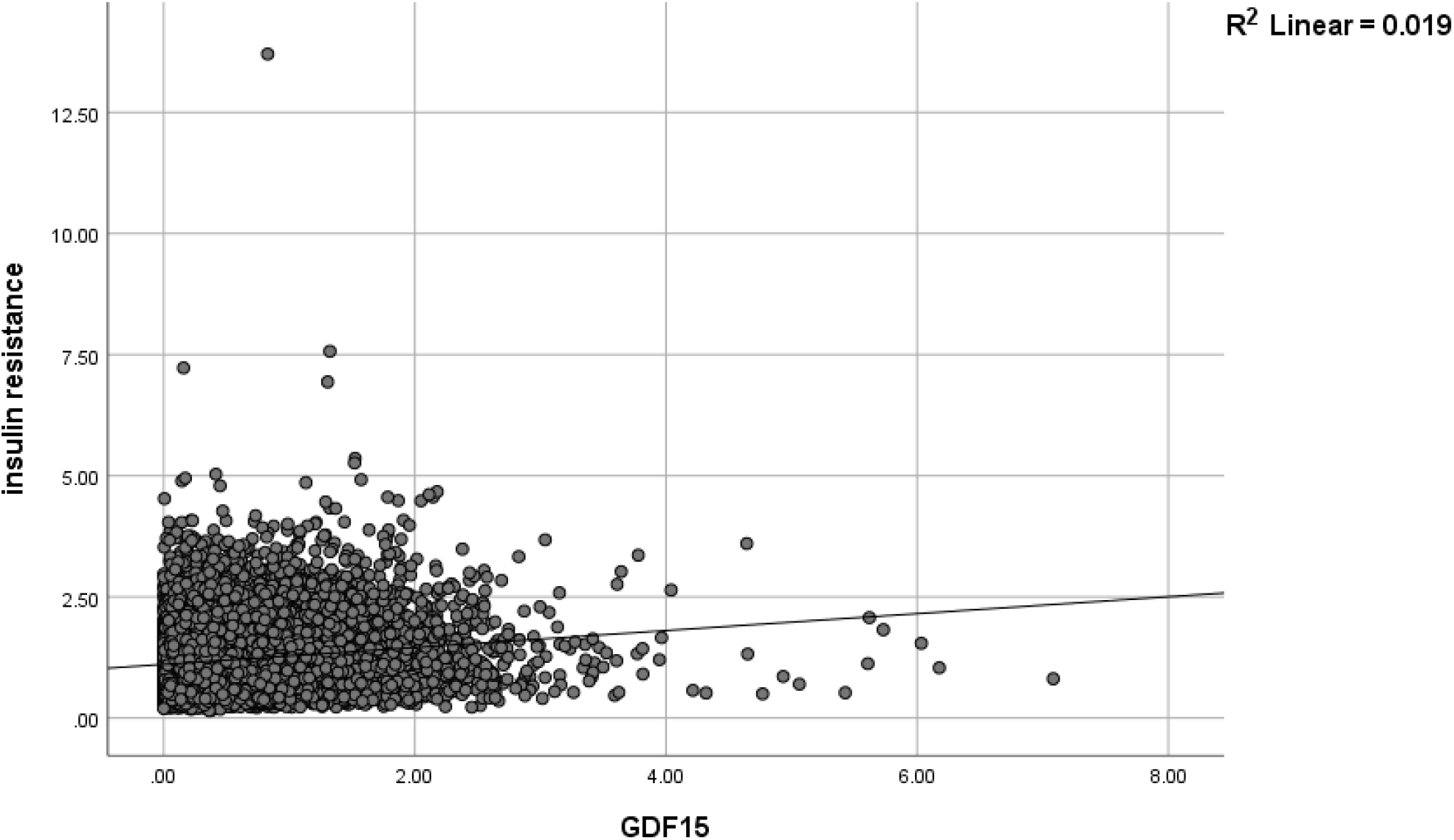
GDF15 versus insulin resistance (triglycerides/HDL-C, 29,811 cases). The correlation is significant (p < 0.001). Individuals with elevated TG/HDL-C have higher insulin resistance.

Figure 3 shows GDF 15 versus hand grip strength right, 25,476 subjects. The correlation is significant (p < 0.001). Individuals with elevated GDF15 have lower hand grip strength.

**Figure 3.**
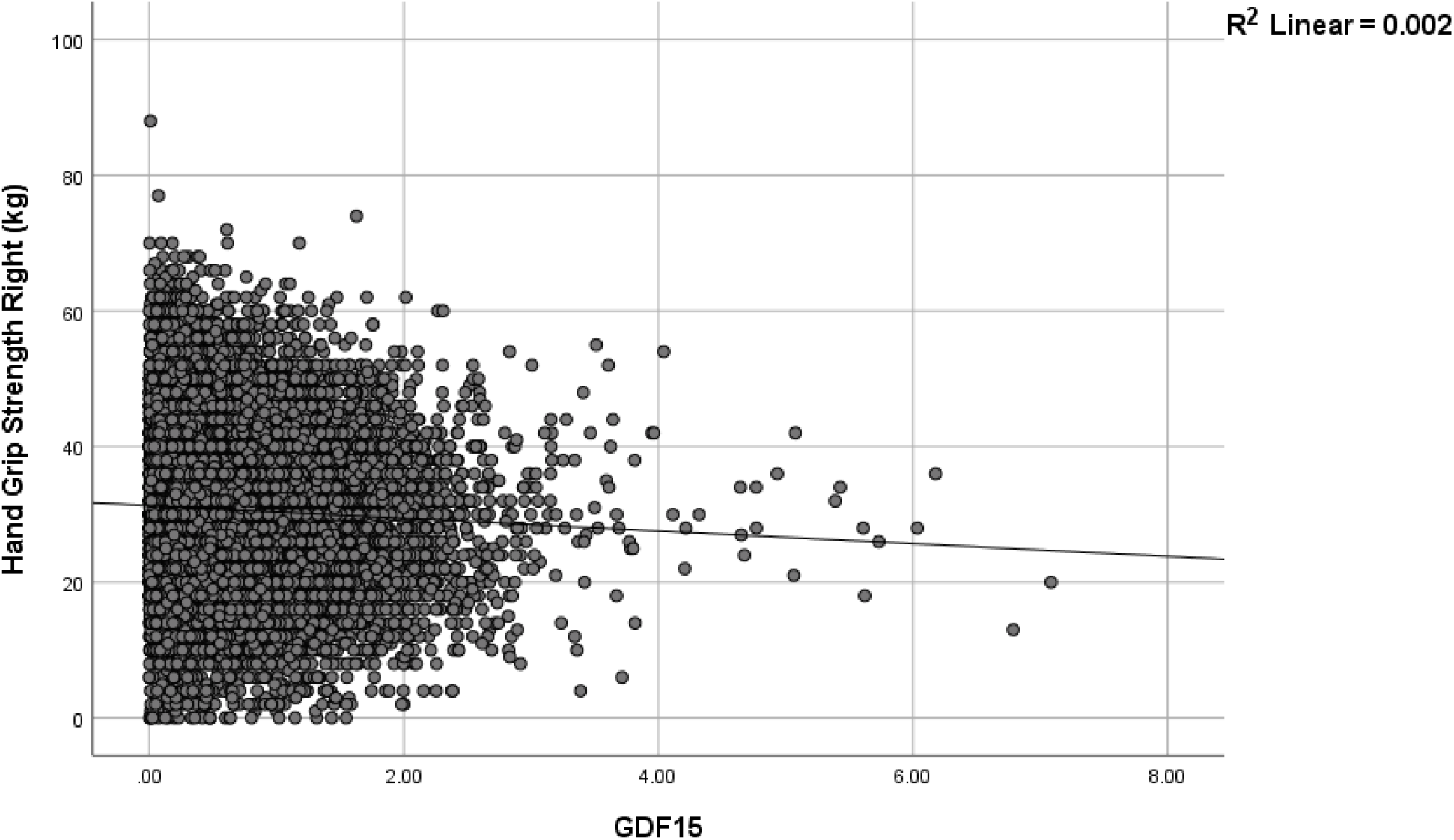
GDF15 versus hand grip strength right, 25,476 subjects. The correlation is significant (p < 0.001). Individuals with elevated GDF15 had lower hand grip strength.

Multivariate linear regression was performed to evaluate the relationship between circulating GDF15 levels and right-hand grip strength, controlling for age and sex. The overall model was significant p < 0.001 and explained 51.3% of the variance in grip strength (R^2^ = 0.513). Higher GDF15 levels were associated with lower grip strength (β = – 0.180, p < 0.001). Age was negatively associated with grip strength (p < 0.001), while male sex was a strong positive predictor (p < 0.001). These results indicate that elevated GDF15 is independently associated with reduced muscle strength.

Figure 4 shows GDF15 versus C Reactive protein, 24,290 subjects. The correlation is significant (p < 0.001). Individuals with elevated GDF15 had increased C reactive protein.

**Figure 4.**
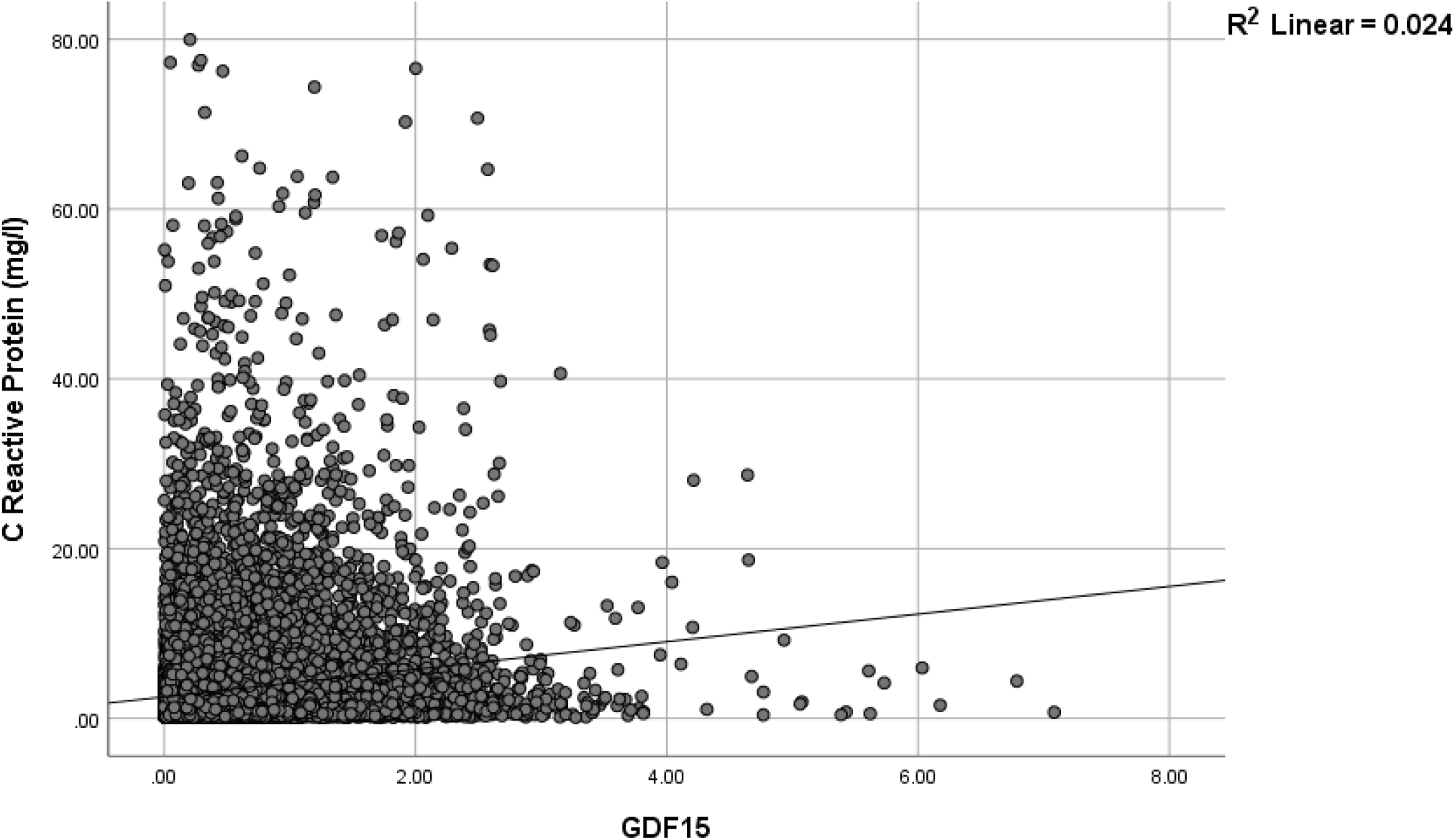
GDF15 versus C Reactive protein, 24,290 subjects. The correlation is significant (p < 0.001). Individuals with elevated GDF15 had increased C reactive protein.

Figure 5 shows GDF15 versus fluid intelligence, 8,264 subjects. The correlation is significant (p < 0.001). Individuals with elevated GDF15 had diminished fluid intelligence.

**Figure 5.**
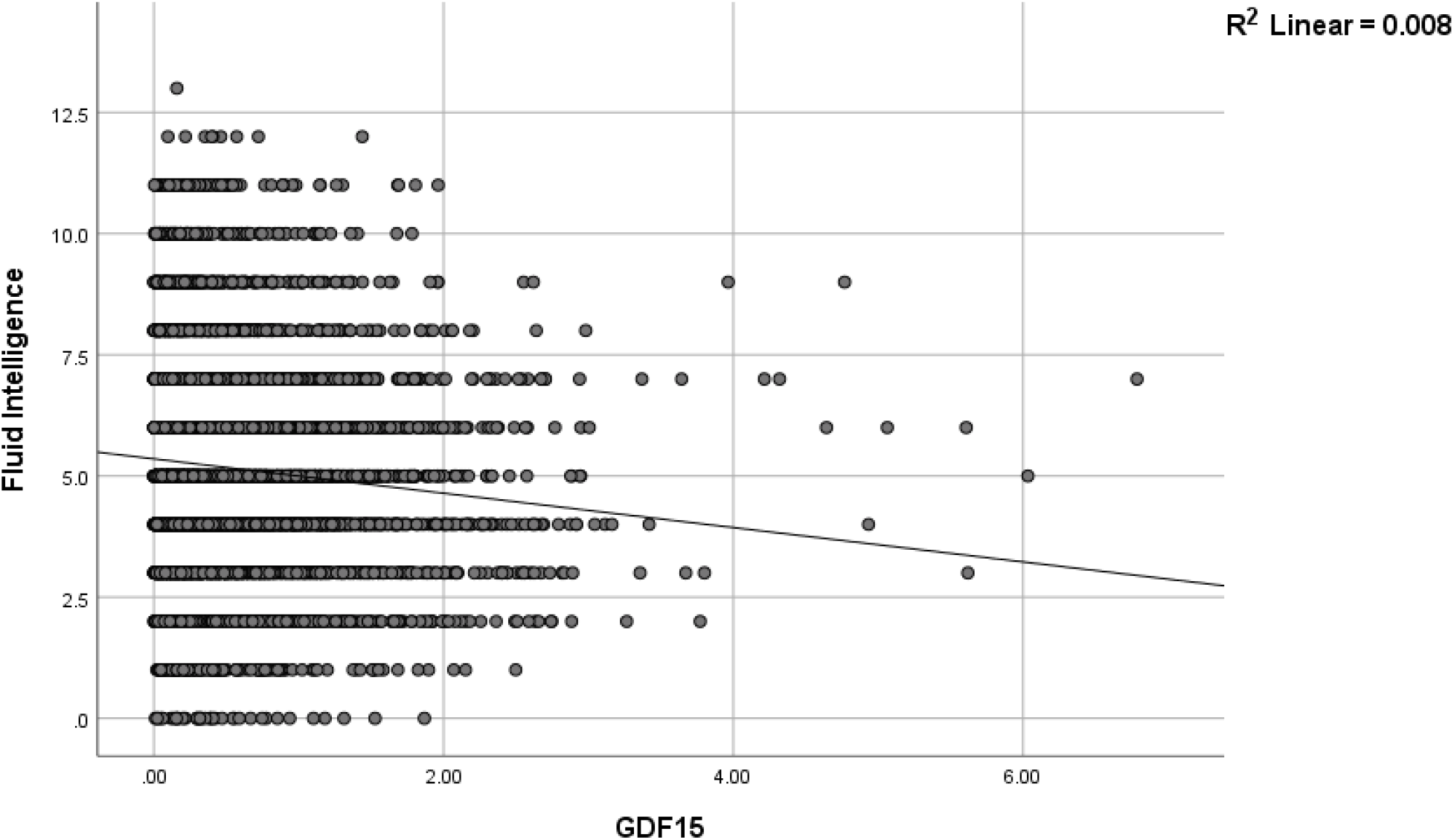
GDF15 versus fluid intelligence, 8,264 subjects. The correlation is significant (p < 0.001). Individuals with elevated GDF15 had lower fluid intelligence.

Multivariate linear regression was performed to evaluate the relationship between circulating GDF15 levels and fluid intelligence, controlling for age, sex, years of education. The overall model was significant p < 0.001 and explained 9.1% of the variance in fluid intelligence (R^2^ = 0.091). Higher GDF15 levels were associated with lower fluid intelligence (β = – 0.058, p < 0.001). Years of education were positively associated with fluid intelligence (β = 0.286, p < 0.001). Age (p = 0.209) and sex (p = 0.234) were insignificantly associated with fluid intelligence. These results indicate that elevated GDF15 is independently associated with diminished fluid intelligence.

Figure 6 shows GDF15 versus Z-standardized telomere length (T/S ratio), 24,674 subjects. The correlation is significant (p < 0.001). Individuals with elevated GDF15 had shorter telomeres.

**Figure 6.**
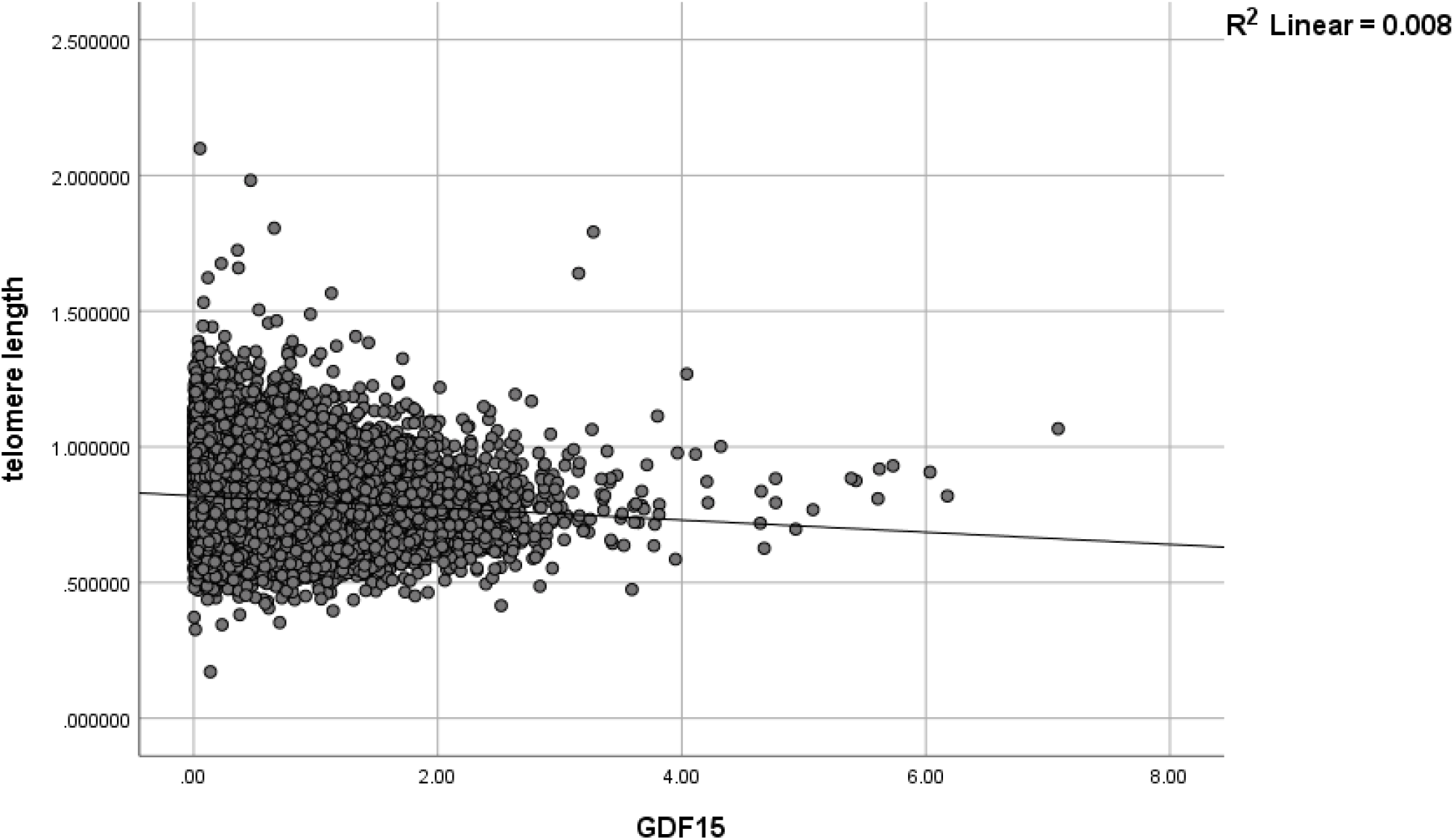
GDF15 versus Z-standardized telomere length (T/S ratio), 24,674 subjects. The correlation is significant (p < 0.001). Individuals with elevated GDF15 had shorter telomeres.

Figure 7 shows GDF15 versus days/week of moderate physical activity 10+ minutes, 23,832 subjects. The correlation is significant (p < 0.001). Individuals with elevated GDF15 had fewer days/week of moderate physical activity.

**Figure 7.**
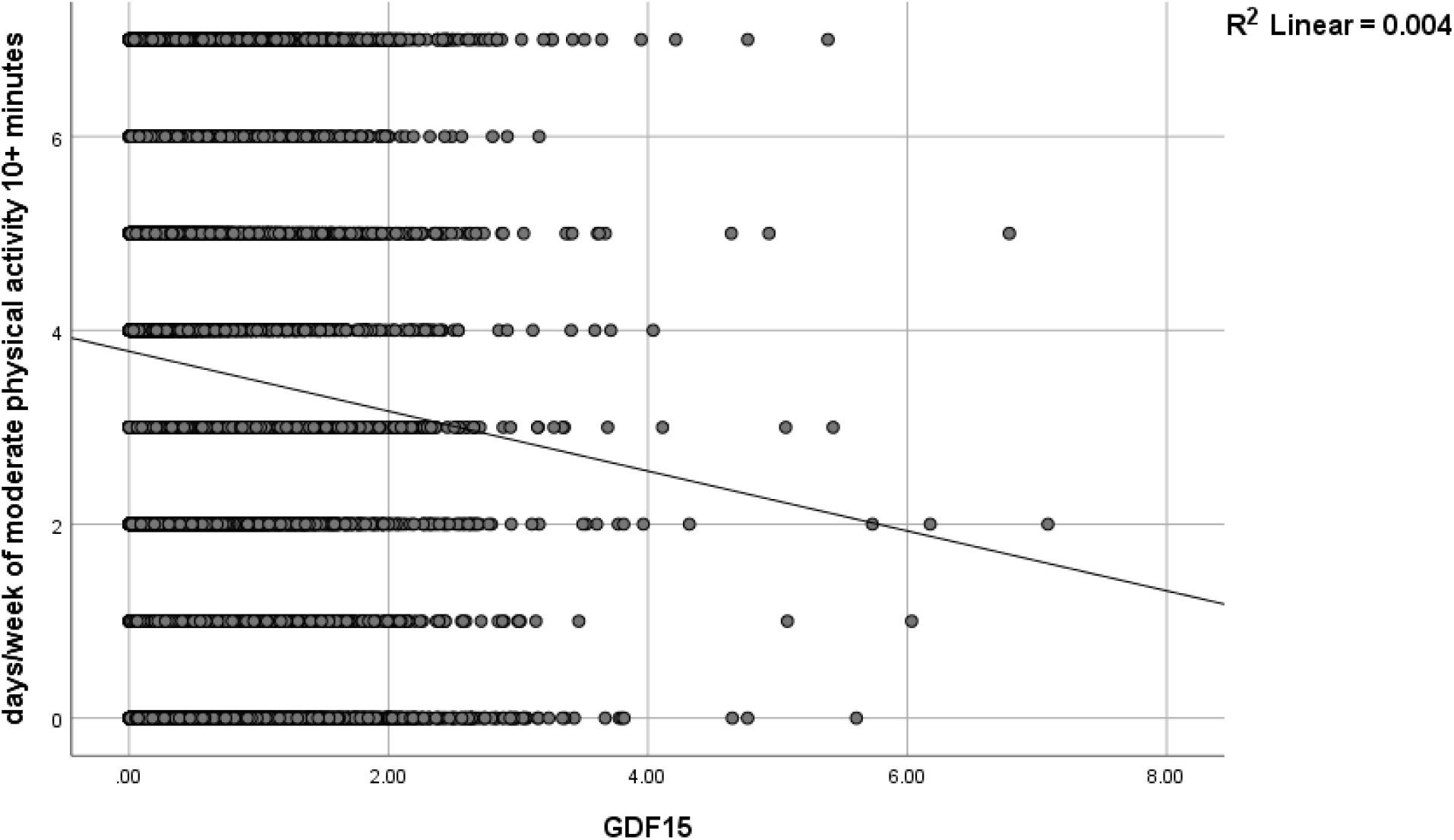
GDF15 versus days/week of moderate physical activity 10+ minutes, 23,832 subjects. The correlation is significant (p < 0.001). Individuals with elevated GDF15 had fewer days/week of moderate physical activity 10+ minutes.

A mediation analysis was conducted to evaluate whether circulating GDF15 mediated the relationship between psychological stress and right-hand grip strength. The overall model predicting GDF15 was significant (R^2^ = 0.0305, p < 0.001), indicating that stress, age, and sex together explained 3.1% of the variance in GDF15. Higher levels of stress were significantly associated with increased GDF15 concentrations (B = 0.084, SE = 0.006, p < 0.001). The model predicting grip strength was also significant (R^2^ = 0.5178, p < 0.001), accounting for 51.8% of the variance. Stress remained directly associated with reduced grip strength (B = –0.856, SE = 0.101, p < 0.001). GDF15 was independently associated with lower grip strength (B = –2.121, SE = 0.099, p < 0.001). Bootstrap analysis with 5,000 samples confirmed a significant indirect effect of stress on grip strength via GDF15 (indirect effect = –0.179, 95% CI –0.213 to –0.147). These findings indicate that GDF15 partially mediates the association between psychological stress and diminished muscle strength.

## Discussion

In this large population-based study, we found that higher circulating GDF15 was consistently associated with multiple indicators of biological ageing, including reduced muscle strength, increased insulin resistance, lower maximum heart rate, shorter telomere length, diminished fluid intelligence, and lower levels of habitual physical activity. These findings align with the brain–body energy-conservation model, which proposes that signals from energetically costly senescent cells prompt the brain to orchestrate systemic metabolic downregulation [1]. Our mediation analysis further showed that GDF15 partially mediated the association between chronic psychological stress and reduced muscle strength, suggesting that stress may accelerate ageing phenotypes in part through upregulation of this cytokine [2, 3]. Together, these results provide empirical support for the hypothesis that GDF15 acts as a circulating signal linking psychosocial and cellular stress to organism-level declines in physiological and cognitive function [4, 5].

Several observations merit emphasis. First, the associations of GDF15 with grip strength and insulin resistance were robust to adjustment for age and sex, underscoring that these relationships are not simply attributable to chronological ageing [4, 5].

Second, the correlation between GDF15 and telomere shortening provides further evidence that this biomarker reflects accumulated cellular stress and senescence [11]. Third, the inverse association between GDF15 and fluid intelligence highlights the possibility that GDF15 may contribute not only to physical frailty but also to cognitive vulnerability [12]. These findings are consistent with previous work linking GDF15 to morbidity and mortality risk [12] and extend prior research by demonstrating that GDF15 is related to self-reported psychological stress and behavioral markers such as physical activity [13, 14].

This study has several limitations. The cross-sectional design precludes determination of causality and temporal sequencing between stress, GDF15 elevation, and functional decline. While our mediation analyses suggest that GDF15 may be a partial mediator, longitudinal studies are needed to establish whether increases in GDF15 precede reductions in muscle strength or cognitive performance [10].

Additionally, although the UK Biobank provides unparalleled sample size and depth of phenotyping, certain measures—including telomere length and fluid intelligence—were available only in subsets of participants, which may limit generalizability [11, 12]. The reliance on self-reported stress exposure is another limitation, as this measure may be subject to recall bias and individual differences in reporting [15]. Finally, unmeasured confounding, such as undiagnosed medical conditions or medication effects, cannot be fully excluded.

In conclusion, our findings support the relevance of GDF15 as a biomarker linking psychological and cellular stress to systemic features of biological ageing. These results lend credence to the brain–body energy-conservation model and highlight the potential value of targeting GDF15-mediated pathways to mitigate stress-related declines in physiological function. Future longitudinal and interventional studies will be critical to clarifying the causal role of GDF15 and to identifying strategies that may preserve resilience in ageing populations. The most promising strategies involve reducing upstream stressors (senescence, mitochondrial dysfunction) and potentially blocking GDF15 signaling at the GFRAL receptor—but these are experimental.

Maintaining muscle mass and metabolic resilience through exercise and nutritional support remains the most evidence-based approach today.

## Data Availability

All data produced in the present work are contained in the manuscript

https://www.ukbiobank.ac.uk/

